# Population-based seroprevalence of SARS-CoV-2 antibodies in a high-altitude setting in Peru

**DOI:** 10.1101/2021.01.17.21249990

**Authors:** Charles Huamaní, Lucio Velásquez, Sonia Montes, Ana Mayanga-Herrera, Antonio Bernabé-Ortiz

## Abstract

**Background:** Little evidence exists about the prevalence of COVID-19 infection at high altitude. We aimed to estimate the population-based seroprevalence of COVID-19 in Cusco at the end of the first wave.

**Methods:** A population-based survey was conducted in September 2020 in three settings in Cusco: (1) Cusco city at 3300 meters above the sea level (m.a.s.l.), (2) the periphery of Cusco (Santiago, San Jerónimo, San Sebastián, and Wanchaq) at 3300 m.a.s.l., and (3) Quillabamba city, located at 1050 m.a.s.l. People aged ≥18 years within a family unit were included. The diagnosis of COVID-19 infection was based on identifying total antibodies (IgM and IgG) anti-SARS-CoV-2 in serum using the Elecsys Anti-SARS-CoV-2 chemiluminescence test.

**Findings:** We enrolled 1924 participants from 712 families. Of the total, 637 participants were COVID-19 seropositive. Seroprevalence was 38·8% (95%CI: 33·4%-44·9%) in Cusco city, 34·9% (95%CI: 30·4%-40·1%) in the periphery of Cusco, and 20·3% (95%CI: 16·2%-25·6%) in Quillabamba. In 141 families (19·8%; 95%CI: 17·0%-22·8%) the whole members were positive to the test. Living with more than three persons in the same house, a positive COVID-19 case at home, and a member who died in the last five months were factors associated with COVID-19 positivity. The smell/taste alteration was the symptom most associated with seropositivity (aOR= 14·27, 95% CI: 8·24-24·70); whereas always wearing a face shield (aOR= 0·62; 95% CI: 0·46-0·84) or a facial mask (aOR= 0·65, 95% CI: 0·47-0·88) reduced that probability.

**Interpretation:** Seroprevalence of COVID-19 in Cusco was high, with significant differences between settings. Wearing masks and face shields were associated with lower rate of infection; however, efforts must be made to sustain them over time since there is still a high proportion of susceptible people.

**Funding:** Fondo Nacional de Desarrollo Científico, Tecnológico y de Innovación Tecnológica (FONDECYT – Perú) and Universidad Andina del Cusco.

## INTRODUCTION

Worldwide, the Coronavirus Disease 19 (COVID-19) pandemic, caused by SARS-CoV-2 virus, has been evaluated in real-time through the official notifications of the affected countries,^1^ which usually come from passive surveillance systems. However, a great proportion of individuals infected by the COVID-19 remains, especially in resource-constrained settings, as it is usually asymptomatic,^2^ and there is a lack of appropriate access to diagnosis in the health care system, both generating a gap in the information for appropriate decisions.^3,4^ To deal with these issues, multiple population-based surveys have been conducted around the world,^5-7^ with divergent results as countries are in different epidemiological scenarios, i.e., beginning or end of the first pandemic wave, urban/rural areas, national/regional representation, or different diagnostic test used (molecular, antibody, or antigen detection tests). Even so, reported prevalence has usually been lower than 20% after the first wave.^6^

Few studies have been carried out in low- and middle-income countries, including Latin American countries,^5-7^ where because different social determinants (e.g. poverty levels, inequities, overcrowding, and a weak health system), a higher COVID-19 prevalence could be expected.^8,9^ Moreover, there are very scarce prevalence studies in high-altitude cities (i.e., those located over 2500 meters above the sea level [m.a.s.l.]). For example, only an ecological study has suggested that the impact of the COVID-19 pandemic would be low because unclear environmental determinants such as atmospheric pressure or radiation.^10^ However, the pandemic progression is more linked to social interaction and adopted preventive measures.^11^

Peru, a country located in South America, has been one of the nations in the top of number of COVID-19 cases worldwide,^1^ and the top three with more deaths in Latin America region with Brazil and Mexico. The Peruvian government response to the pandemic was prompt, including mandatory social distancing measures and re-focusing most health system resources to address the pandemic. Peru is a very heterogeneous country with a variable geography, thus, although the first case of COVID-19 was detected in March 2020, some high-altitude cities like Cusco experienced the first wave between July and September.^12^ At the end of this period, we conducted a population-based seroprevalence survey in different areas of Cusco, and evaluated some determinants associated with the spread of the COVID-19 infection.

## MATERIALS AND METHODS

### Study design

A population-based cross-sectional study was conducted between September 12 and 27, 2020; as part of a longitudinal cohort study to determine the prevalence and incidence of COVID-19 infection and propagation factors in high-altitude residents (Cusco, 3300 m.a.s.l.).

### Study locations

The region of Cusco comprises thirteen provinces, each having a different number of cities (Figure 1). Cities offering variability of scenarios were chosen due to their proportion of urban/rural areas, altitude, and population. Three different study settings were selected as follows:

**Figure 1.**
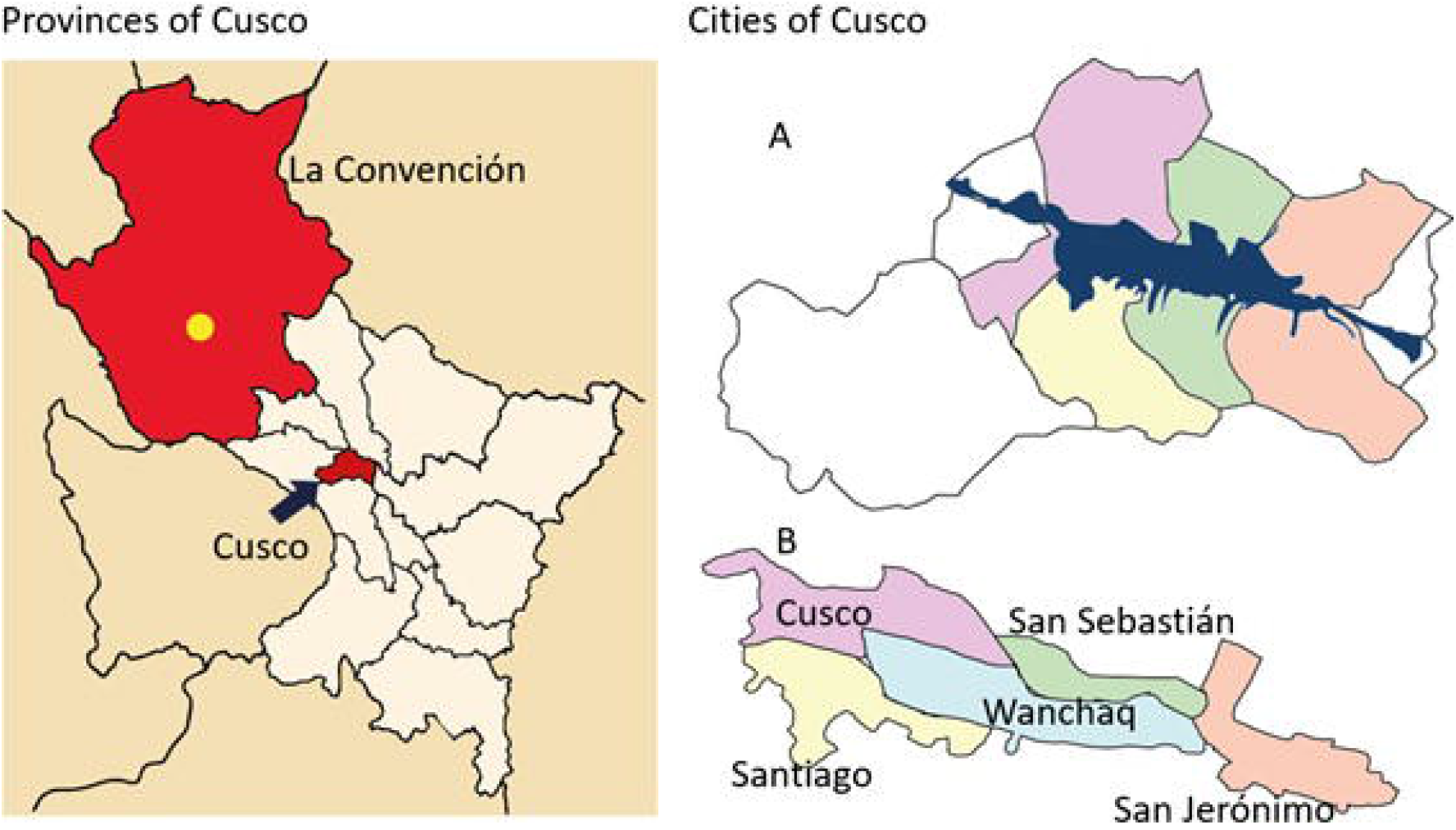
Map of Cusco, provinces, and cities. Cusco has thirteen provinces (left picture). The capital of La Convención Province is Quillabamba (yellow dot), located at 1050 m.a.s.l. The Cusco Province (blue arrow), at 3300 m.a.s.l., is inhabited in only one sector located in the valleys (blue mark in A), which include the five cities where the research was carried out (B).

- *Cusco city, Province of Cusco*, with a high demographic density (approximately 1000 inhabitants/km^2^), population (125 thousand inhabitants), and located at 3330 m.a.s.l. Based on the information available about the behavior of the COVID-19 pandemic in the region,^12^ it would represent the worst transmission scenario.
*- Periphery of Cusco, Province of Cusco*, including different cities around the Cusco city (Santiago, San Sebastián, San Jerónimo, and Wanchaq), where approximately 320,000 inhabitants in total live. Although these cities are at the same altitude as Cusco city, they are located on the historic center’s periphery.
- *Quillabamba City, Province of La Convención*, located at the lowest altitude in the department of Cusco (1050 m.a.s.l.) with an approximate population of 30,000 inhabitants, and geographically distant (approximately 6 hours by road, not accessible by any other way), with a low population density (<10 inhabitants/km^2^). This city is part of the Peruvian jungle and, as a result, the climate and geography are different from that of Cusco city. Despite of that, during the first wave, La Convención was the second Province of Cusco with the most reported cases.^12^

### Participants and sampling

People aged ≥18 years old, who voluntarily agreed to participate in the study, signed their informed consent, accepted the telephone and serological follow-up, usual resident of the study area (≥6 months), and with the ability to understand the procedures were included. We did not exclude people with acute symptoms or who had already given positive results to previous tests.

A two-stage probability sampling approach was carried out: by clusters of groups of blocks and by households. Maps by each of the cities were used as sampling frame. The primary sampling unit was defined as the cluster comprised by a block or group of blocks with approximately 40 households. In the Cusco city, 98 clusters were created, whereas these numbers were 495 and 74 for the periphery of Cusco and Quillabamba, respectively. Of them, 10, 16 and 8 clusters were randomly chosen in each of the settings, respectively. Within each cluster, households were selected randomly. In each selected household, all the members who met the eligibility criteria were included until the intended sample size was achieved.

The sample size calculation was based on an expected population prevalence of at least 5%, precision of 2·5%, a confidence level of 95%, and a design effect of 2·0. Based on these estimates, the required sample size in a conservative setting was 1752 participants for the three settings. With this sample size, we had a power over 80% to detect a difference in the prevalence of COVID-19 infection of at least 5% (e.g., 5% vs. 10%) between the groups of interest (study setting and gender). Up to 1800 participants were enrolled for the three settings, considering losses to follow-up, and split into 600 participants for Cusco city, 800 for the periphery of Cusco, and 400 for Quillabamba.

### Procedures

The collection of information and biological samples was carried out during three weekends in September 2020 (12, 13, 19, 20, 26 and 27) taking advantage of the period of lockdown in the region. Data and sample collection were prioritized during Sundays to guarantee the presence of the largest number of family members. The field staff visited each household to contact potential participants, assess eligibility, invite them to participate in the study, apply the informed consent, questionnaires, and finally, take blood samples.

The questionnaire evaluated sociodemographic characteristics (age, gender, etc.), risk history (self-report of hypertension, type 2 diabetes, asthma, etc.), weight and height (self-reported), number of people living together, past COVID-19 case at home, member who died in the last five months, among others. We included questions about the symptoms developed in the last three months, previous COVID-19 tests, and protective behaviors used or applied during quarantine (use of a mask, alcohol, gloves, etc.).

Once the questionnaire was completed, a blood sample of 3·5 mL of whole blood was taken in serum separating tubes. Each tube was coded and stored for transportation. Sample conservation criteria for several days were according to the World Health Organization guidelines.^13^

Chemiluminescence tests were used for serological detection of antibodies against SARS-CoV-2 Elecsys from the ROCHE laboratory. This test is based on a sandwich-type immunoassay, where the recombinant protein N of SARS-CoV-2 is the target detected by possible antibodies present in the serum sample.^14^ The test detects total antibodies simultaneously, without differentiating between IgM or IgG. This test has a specificity of 99·5%, and whether the test is performed after 14 days of having a positive result by PCR, the sensitivity may reach up to 99.8%.^14,15^

### Statistical analysis

For data analysis, STATA 16 for Windows (StataCorp, CollegeStation, TX, US) was utilized. The description of the study population was carried out according to the characteristics of interest. The prevalence of COVID-19 infection was estimated, taking into account the sampling techniques used. Prevalence estimates were calculated using Poisson distribution models adjusting for clusters at household level and with robust variance. To facilitate a comparison of our results with other studies, a mixed-effect multilevel logistic regression model was used to identify the factors associated with positivity for COVID-19, obtaining adjusted odds ratios (aOR) by study settings, gender, and age group (<40, 40-59, and 60 + years of age), considering clusters at the household level. All estimates are presented with 95% confidence intervals (95%CI).

### Ethics

The study was approved by the Ethics Committee of the Universidad Científica del Sur (code 051-2020-PRO99). Written informed consent was used, and although personal identifiers were collected, this was done to guarantee appropriate follow-up and deliver test results. All the results were delivered directly to study participants. To reduce the risks of contagion by COVID-19 during the execution of the research project, all the interviewers followed the protocols for handling COVID-19 patients and providing masks to study participants. The participants had access to help lines, where trained physicians answered their questions and provided guidance concerning COVID-19.

### Patient and Public Involvement

Patients or the public were not involved in the design, or conduct, or reporting. The public were involved in the dissemination plans of our research. Therefore, our results were broadcast on local television stations in Cusco, newspapers and web pages.

## RESULTS

### Characteristics of the study population

A total of 712 families (range: 1 to 11 members included) were enrolled in the present study, with a total of 1924 participants being evaluated. Of them, 408 (21·2%) were from Quillabamba, 640 (33·3%) were from Cusco city, and 876 (45·5%) were from the periphery of Cusco. The average age was 42·5 (SD: 16·5), and 1096 (57·1%) were women, without these characteristics being different among the different study settings. Some differences between the settings, however, were observed; thus, self-reported obesity was more frequent (26·5%) in Quillabamba compared to Cusco city (13·2%) or the periphery of Cusco (18·3%, p-value <0·001). The characteristics of the study population, according to the study settings, are shown in Table 1.

**Table 1:** Population characteristics according to the study settings

|  | **TOTAL**<br>(n = 1924) | Cusco city<br>(n = 640) | Periphery of Cusco<br>(n = 876) | Quillabamba<br>(n = 408) | p-value |
| --- | --- | --- | --- | --- | --- |
| **Gender** |  |  |  |  |  |
| Female | 1096 (57.1%) | 359 (56.3%) | 500 (57.1%) | 237 (58.2%) | 0.822 |
| Age: Media (S.D.) | 42.47 (16.5) | 42.8 (16.2) | 42.5 (16.8) | 42.81 (16.5) | 0.884 |
| **Age group (years)** |  |  |  |  |  |
| <40 | 876 (46.5%) | 298 (47.3%) | 403 (46.8%) | 175 (44.4%) | 0.843 |
| 40-59 | 702 (37.2%) | 235 (37.3%) | 313 (36.3%) | 154 (39.1%) |  |
| > 60 | 308 (16.3%) | 97 (15.4%) | 146 (16.9%) | 65 (16.5%) |  |
| People living in the same house: Media (SD) | 5.98 (3.43) | 6.24 (3.40) | 6.16 (3.58) | 5.04 (2.92) | <0.001 |
| Previous COVID-19 test | 758 (39.7%) | 242 (38.1%) | 328 (37.7%) | 188 (46.7%) | 0.006 |
| Asymptomatic during pandemic | 772 (40.2%) | 236 (36.9%) | 300 (34.4%) | 236 (57.9%) | <0.001 |
| **Personal history** |  |  |  |  |  |
| Obese | 332 (18.4%) | 86 (13.2%) | 152 (18.3%) | 94 (26.5%) | <0.001 |
| Hypertension | 128 (6.7%) | 43 (6.7%) | 55 (6.3%) | 30 (7.4%) | 0.763 |
| Type 2 diabetes | 85 (4.4%) | 19 (3.0%) | 37 (4.2%) | 29 (7.1%) | 0.006 |
| Renal disease | 19 (1.0%) | 5 (0.8%) | 12 (1.4%) | 2 (0.5%) | 0.271 |
| Asthma | 29 (1.5%) | 10 (1.6%) | 13 (1.5%) | 6 (1.5%) | 0.990 |

Overall, 39·7% (95%CI: 36·8%-42·9%) of participants indicated at least one previous test to detect COVID-19. The factors associated with decreasing the probability of doing it were being asymptomatic (aOR= 0·68, 95% CI: 0·55-0·85) or being a woman (aOR = 0·75, 95% CI: 0·63-0·90); whilst, higher education (> 12 years) increased the probability of taking the test (aOR = 2·15, 95% CI: 1·43-3·22).

From 606 families that reported information, 46 (7·6%) stated that at least one member deceased in the five months before the survey, and from these, 60·5% attributed the cause of death to COVID-19 infection.

### Prevalence of COVID-19 infection

A total of 637 participants were reactive to the screening test, which defines an adjusted prevalence of 33·1% (95%CI: 30·1%-36·4%). This prevalence varied according to the study settings: 20·3% (95%CI: 16·2%-25·6%) in Quillabamba, 38·8% (95% CI: 33·4%-44·9%) in Cusco city, and 34·9% (95%CI: 30·4%-40·1%) in the periphery of Cusco (Table 2).

**Table 2:** Prevalence of COVID-19 according to individual/family characteristics

|  | Prevalence | Adjusted OR |
| --- | --- | --- |
| *Global prevalence* | 33.1% (30.1 - 36.4%) |  |
| *Gender* |  |  |
| Male | 31.1% (27.3 -35.2%) | 1 |
| Female | 34.6% (31.2 -38.3%) | 1.17 (0.98-1.41) |
| *Study setting* |  |  |
| Cusco city | 38.8% (33.4 -44.9%) | **2.39 (1.63-3.49)** |
| Periphery of Cusco | 34.9% (30.4 -40.1%) | **2.06 (1.44-2.95)** |
| Quillabamba | 20.3% (16.2 -25.6%) | **1** |
| *Age group (years)* |  |  |
| <40 | 34.8% (31.0 -39.1%) | 1 |
| 40-59 | 35.8% (31.8 -40.2%) | 1.05 (0.85-1.29) |
| > 60 | 24.7% (19.7 -30.9%) | **0.62 (0.46-0.85)** |
| *Education level (years)* |  |  |
| <7 | 34.2% (26.9-43.3%) | 1 |
| 7 -11 | 39.2% (34.4-44.7%) | 1.17 (0.78-1.75) |
| > 12 | 30.3% (26.9-34.0%) | 0.70 (0.47-1.06) |
| *BMI* |  |  |
| Normal | 34.1% (30.0-38.6%) | 1 |
| Overweight | 31.1% (27.1-35.7%) | 0.94 (0.73-1.20) |
| Obese | 35.5% (30.2-41.8%) | 1.28 (0.92-1.78) |
| *Number of people living together* |  |  |
| 1 - 2 | 20.5% (14.4-29.1%) | **1** |
| 3 - 5 | 31.8% (27.6-36.7%) | **1.71 (1.04-2.81)** |
| 6 -10 | 37.5% (31.9-44.0%) | **2.10 (1.25-3.52)** |
| > 11 | 38.7% (28.1-53.2%) | **2.18 (1.09-4.33)** |
| *Self-report of COVID-19 case at home* |  |  |
| No | 26.2% (22.9-29.9%) | **1** |
| Yes | 50.2% (44.4-56.7%) | **2.92 (2.16-3.96)** |
| *Death at home (any cause)* |  |  |
| No | 31.9% (28.7-35.3%) | **1** |
| Yes | 50.8% (38.4-67.2%) | **2.06 (1.11-3.83)** |
| *Previous COVID-19 test* |  |  |
| No | 28.7% (25.3-32.6%) | **1** |
| Yes | 39.9% (35.7-44.7%) | **1.82 (1.44-2.29)** |

Models adjusted by age, gender, and study setting

Of the 712 families evaluated, 318 (44·6%; 95% CI: 41·0%-48·3%) had at least one infected member, and 141 (19·8%; 95% CI: 17·0%-22·8%) families had the whole members positive to the test.

### Factors associated with positivity for COVID-19

Characteristics such as gender, education level, BMI category, or personal history of diseases were not associated with changes in the probability of being positive for COVID-19 (p>0·05). The prevalence of COVID-19 infection was lower in those aged ≥60 years (26·2%; 95%CI: 20·7%-33·1%). Other factors associated with COVID-19 infection were living with 3 or more people, a family member with previous COVID-19 infection, and have a death in the household in the last five months (Table 3).

**Table 3:** Factors associated with positivity for COVID-19

|  | Negatives<br>**1284 (100%)** | Positives<br>**636 (100%)** | Adjusted OR |
| --- | --- | --- | --- |
| *PERSONAL HISTORY* |  |  |  |
| *HBP* | 101 (7.85%) | 27 (4.24%) | 0.61 (0.37-1.00) |
| *Diabetes* | 64 (4.98%) | 21 (3.30%) | 0.82 (0.48-1.40) |
| *Asthma* | 23 (1.79%) | 6 (0.94%) | 0.52 (0.20-1.35) |
| *Cardiac disease* | 11 (0.87%) | 1 (0.16%) | 0.22 (0.03-1.78) |
| *Renal disease* | 12 (0.93%) | 7 (1.10%) | 1.18 (0.46-3.02) |
| *Cancer* | 9 (0.70%) | 1 (0.16%) | 0.21 (0.03-1.64) |
| *DEVELOPED SOME OF THESE SYMPTOMS IN THE LAST 3 MONTHS* |  |  |  |
| *No symptoms* | 584 (45.5%) | 188 (29.6%) | **0.57 (0.45-0.73)** |
| *Smell / taste alteration* | 18 (1.4%) | 118 (18.6%) | **14.27 (8.24-24.7)** |
| *Respiratory distress* | 23 (1.8%) | 65 (10.2%) | **5.72 (3.21-10.18)** |
| *Fever* | 108 (8.4%) | 138 (21.7%) | **2.79 (2.06-3.77)** |
| *General discomfort* | 136 (10.6%) | 163 (25.6%) | **2.69 (2.01-3.60)** |
| *Cough* | 174 (13.5%) | 178 (27.9%) | **2.45 (1.92-3.12)** |
| *Muscle pain* | 146 (11.4%) | 149 (23.4%) | **2.18 (1.64-2.90)** |
| *Nasal congestion* | 105 (8.2%) | 102 (16.0%) | **1.94 (1.39-2.70)** |
| *Back pain* | 216 (16.8%) | 182 (28.6%) | **1.88 (1.43-2.46)** |
| *Diarrhea* | 96 (7.5%) | 82 (12.9%) | **1.77 (1.24-2.51)** |
| *Headache* | 412 (32.0%) | 275 (43.2%) | **1.41 (1.12-1.78)** |
| *Sore throat* | 272 (21.2%) | 167 (26.2%) | 1.17 (0.90-1.52) |
| *BEHAVIORS* |  |  |  |
| *Always wear a face shield* | 338 (27.6%) | 139 (23.0%) | **0.62 (0.46-0.84)** |
| *Always wear a face mask* | 1085 (85.8%) | 511 (81.5%) | **0.65 (0.47-0.88)** |
| *Always use alcohol* | 1039 (82.7%) | 486 (77.5%) | **0.67 (0.49-0.89)** |
| *Always wash hands* | 1036 (83.7%) | 484 (78.3%) | 0.74 (0.54-1.00) |
| *Always stay more than 1 meter away from other people* | 945 (75.3%) | 443 (70.8%) | 0.78 (0.61-1.00) |
| *Always wear gloves* | 69 (5.9%) | 31 (5.2%) | 0.72 (0.45-1.17) |
| Models adjusted by age, gender, and study setting |  |  |  |

Overall, 40·2% (95% CI: 37·3%-43·4%) of participants had no symptoms in the three months prior to the interview; however, 24·4% (95% CI 20·7%-28·6%) of them were positive for COVID-19. Among those who developed any symptom, 39·0% (95% CI: 35·2%-43·2%) were positive for COVID-19. Having no symptoms was associated with a 43% reduction in the probability of having COVID-19 (aOR =0·57, 95%CI: 0·44-0·73); however, some symptoms increased the probability of having COVID-19, such as smell/taste alteration (aOR = 14·27, 95CI% 8·24-24·7) and respiratory distress (aOR = 5·72, 95%CI: 3·21-10·18).

Finally, 1473 (92·2%) reported accomplishing with some protective behaviors; the use of at least one of them decreased the probability of having COVID-19 by 48% (aOR= 0·52, 95% CI: 0·33-0·82). Specifically, always wearing a face shield decreased by 38% (aOR = 0·62, 95% CI: 0·46-0·84) the probability of being positive for COVID-19, whereas the use of face masks decreased it by 35% (aOR= 0·65, 95% CI: 0·47-0·88), and the use of alcohol by 33% (aOR =0·67, 95% CI 0·49-0·89).

## DISCUSSION

Our results indicate that, on average, a third of the population of Cusco had antibodies against SARS-CoV-2 virus, expanding our knowledge about the epidemiology of COVID-19 pandemic in high-altitude settings, a very high result compared to other seroprevalence studies in different settings and countries. These results may be appropriately contextualized to understand the varying spread and transmission of the epidemic.^3,16^ In Peru, a mandatory social lockdown, suspension of tourism, and internal migration were quickly adopted after the detection of the first COVID-19 case;^17^ therefore, the first wave was presented relatively late compared to other countries in the region. Cusco reported the first positive case in March 2020; but it was not until June that a sustained increase in cases was identified, reaching the peak in August, and a decreased occurred since September.^12^ Thus, our results were collected immediately after the end of the first wave in Cusco, and results should be compared with studies in a similar epidemiological situation.

Prevalence of COVID-19 infection may differ depending upon the pandemic stage. A nationwide study in Spain reported a prevalence of 5%^18^ with markedly geographical variation (up to 10% in Madrid); however, a study in a region of Brazil, reported a population-based prevalence of 40%.^19^ In Peru, some seroprevalence studies have already been reported; for example, during the last days of June 2020, the prevalence in Lima was estimated in 25·3%,^20^ while in Cusco was 2·6%.^21^ However, Lima had the peak of infections in June 2020 compared to August in Cusco. Other Peruvian cities have also reported high prevalence at the end of their first wave, such as Lambayeque (located on the northern coast of Peru and strongly affected) where the prevalence was 30% by September, and it was 71% in Iquitos (located in the Peruvian jungle), the highest reported in the country.^22^ These studies used lateral immunochromatography tests (rapid tests) as a diagnostic method, which have obtained a diagnostic yield of less than 50% sensitivity in a field validation in Peru.^23^ We used, however, chemiluminescence tests with a diagnostic yield greater than 95% to adequately report our estimates.

In addition, our results show a marked difference in seroprevalence rates between cities in the same region. While in Cusco region, on average, we have a prevalence of 33%, this estimate in Cusco city almost doubled that of Quillabamba (38% vs. 20%). The national seroprevalence study, carried out in April 2020, in Spain,^18^ has already shown that cities with>100,000 inhabitants have higher prevalence, probably due to more significant social interaction, as could happen in our study, where the Cusco city, with high demographic density, and the periphery of that city, were markedly affected.

Contrary to the postulates presented in ecological studies that evaluated contagion in high-altitude cities,^10,24^ the prevalence in our study was inverse to the altitude of the city, but directly correlated with population density. Altitude does not appear to be an associated factor, though it will not be easily determined. Our study shows that in an urbanized city with a high population density, the prevalence is high regardless of the altitude at which they are located. These factors would condition the prevalence of a disease;^11^ ecological studies do not usually include analyses of these confounding variables.

Our results are consistent regarding some factors associated with a greater probability of positivity to COVID-19, for example, the symptoms described such as anosmia or respiratory distress, in the context of the pandemic, have a strong disease predictive association.^25^ In recent systematic reviews and meta-analyses, anosmia has been similarly associated with COVID-19 infection with an OR between 11 and 14,^26,27^ like in our study. These findings could be overestimated as the prevalence of our outcome was high (> 20%), but when correcting the analyzes using Prevalence Ratios (PR) for smell/taste alteration, the PR was 2·74 (2·41-3·12), a significant association but with lower point estimates.

At the beginning of the pandemic, the first publications focused on contagion processes between contiguous people, such as clusters of families.^28,29^ These reports indicated that transmission between members of a family was not total, despite the existence of contact between the family members. The secondary attack rate among members of a family was around 36%,^30^ however, in population seroprevalence studies, intra-domiciliary transmission has not been studied. In our study, if one family member was positive, in 44% of the cases all family members were positive. This result may be more relevant for the purposes of adjusting the mathematical models of intra-household transmission.

Chronic diseases, such as hypertension, type 2 diabetes or obesity have already been described as predictors of poor prognosis of COVID-19,^31^ therefore in Peru, the isolation of people with these diseases was recommended. Our study did not evidence that people with these antecedents had a different probability of contagion. In contrast, the older age groups (> 60 years) do present a lower risk, perhaps due to the lower exposure as they do not have the same needs to leave home or because the other members of their family protected them.

The actions of social distancing, wearing masks, and protection, in general, are included in several recommendations to reduce the spread of the infection, although there were discrepancies in their use (limit it only to symptomatic people, people at risk, among others).^32^ The quantitative reduction value in contagion possibilities was evaluated in multiple studies, which resulted in a meta-analysis with positive results in favor of these measures.^33^ We identified a protective factor between 30% and 35% in practicing these measures, which leads us to continue recommending them. Although there is a decrease in risk, we could not identify some protective factors such as social distancing or hand washing, due to insufficient statistical power.

Since these measures are effective, inexpensive and easy to implement, it would be expected to be of general use. A study carried out in China during the pandemic early phase found that at least 84% of the population complied with some protection measure,^34^ in our study at the first wave end it was observed that 92% of participants reported always followed some of the measures evaluated. Although the use was better accepted at the beginning of the pandemic, after the first wave, a decrease in use was expected, especially since Peru does not have a tradition in the widespread and constant use of masks or other protective measures.

Our study is not representative for the whole Cusco region because of access to cities and distance among them was not easy in lockdown times. However, the representativeness of the cities with the largest population size and most affected was prioritized to facilitate decision-making in public health in the region, knowing that the prevalence is also lower in the cities with smaller population size. Additionally, despite the indication of lockdown at the beginning of our study, the population had to work outside their homes; for this reason, the sample collection was mainly on Sundays to guarantee representativeness of the population. Finally, lack of power can be an issue as some associations were not significant, although these showed a suggestive trend.

In conclusion, at the end of the first wave, the seroprevalence found in Cusco, a high-altitude region, was high, and the altitude seems not to influence estimates, as done by other factors such as population density or population size. The factors associated with a lower probability of having COVID-19 have been widely recommended (wearing a mask, use of alcohol, hand washing), but efforts must be made to sustain them over time since there is still a high proportion of susceptible people.

## Data Availability

The data are not publicly available.

## Acknowledgements

We thank the personnel who participated in the collection of samples, transportation, and digitization; and to the patients, who facilitated the execution of the work.

